# Model to Describe Fast Shutoff of CoVID-19 Pandemic Spread

**DOI:** 10.1101/2020.08.07.20169904

**Authors:** Genghmun Eng

## Abstract

Early CoVID-19 growth obeys: 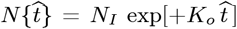, with *K_o_* = [(ln 2)/(*t_dbl_*)], where *t_dbl_* is the pandemic growth *doubling time*. Given 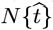, the daily number of new CoVID-19 cases is 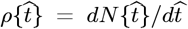. Implementing society-wide *Social Distancing* increases the *t_dbl_ doubling time*, and a linear function of time for *t_dbl_* was used in our *Initial Model*:

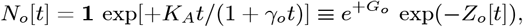

to describe these changes, with *G_o_* = [*K_A_*/*γ_o_*]. However, this equation could not easily model some quickly decreasing *ρ*[*t*] cases, indicating that a second *Social Distancing* process was involved. This second process is most evident in the initial CoVID-19 data from *China, South Korea*, and *Italy*. The *Italy* data is analyzed here in detail as representative of this second process. Modifying *Z_o_*[*t*] to allow exponential cutoffs:

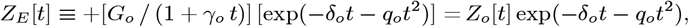

provides a new *Enhanced Initial Model (EIM)*, which significantly improves datafits, where 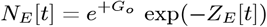. Since large variations are present in *ρ_data_* [*t*], these models were generalized into an orthogonal function series, to provide additional data fitting parameters:

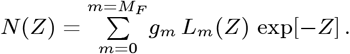

Its first term can give *N_o_*[*t*] or *N_E_* [*t*], for *Z*[*t*] → *Z_o_*[*t*] or Z[t] → *Z_e_*[*t*]. The *L_m_*(*Z*) are *Laguerre Polynomials*, with *L*_0_(*Z*) = 1, and {*g_m_*; *m* = *0,M_F_*} are constants derived from each dataset. When *ρ*[*t*] = *dN*[*t*]/*dt* gradually decreases, using *Z_o_*[*t*] provided good datafits at small *M_F_* values, but was inadequate if *ρ*[*t*] decreased faster. For those cases, *Z_E_*[*t*] was used in the above *N*(*Z*) series to give the most general *Enhanced Orthogonal Function [EOF]* model developed here. Even with *M_F_* = 0, *q_o_* = 0, this *EOF* model fit the *Italy* CoVID-19 data for *ρ*[*t*] ≡ *dN*[*t*]/*dt* fairly well.

When the *ρ*[*t*] post-peak behavior is not Gaussian, then *Z_E_*[*t*] with *δ_o_* ≠ 0, *q_o_* = 0; which we call *Z_A_*[*t*], is also likely to be a sufficient extension of the *Z_o_*[*t*] model. The *EOF* model also can model a gradually decreasing *ρ*[*t*] tail using small {*δ_o_*, *q_o_*} values [with *6 Figures*].

## 1 Introduction

Let 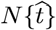 be the total number of CoVID-19 cases in any given locality, with 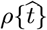 being the predicted number of daily new CoVID-19 cases, so that:

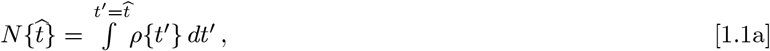

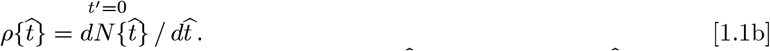

Early CoVID-19 growth often obeys 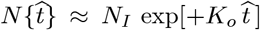, with *K_o_* = [(ln2)/*t_dbl_*, where *t_dbl_* is the pandemic *doubling time*. The start of society-wide *Social Distancing* at 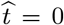 can gradually lengthen *t_dbl_* for 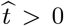. The 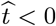 exponential growth phase is not applicable for estimating *Social Distancing* effects. For 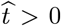, an *Initial Model* for CoVID-19 pandemic shutoff was first developed**^1^** using a linear function of time to describe the *t_dbl_* changes:

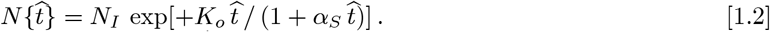

Given measured 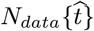, the data end-points {*N_I_*, *N_F_*} help to set {*K_o_*, *α_S_*}. An *Orthogonal Function Model [OFM]* was developed next**^2^**, with Eq. [1.2] as the first term of the orthogonal function series. Each new *OFM* term provides another fitting parameter, to progressively better match 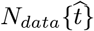 and 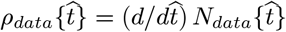.

The *OFM* improves on the *Initial Model*, and it works best with gradually decreasing 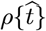 [“Slow *Shutoff*”]. In contrast, when 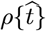 decreased quickly [“Fast *Shutoff*”], the *Initial Model* was not a good datafit, and a few-term *OFM* series only gave small improvements. This result indicates there is an inherent limit to what the gradually changing *t_dbl_ doubling time* of Eq. [1.2] can model.

For these cases, typified by CoVID-19 pandemic evolution in Italy, data often showed a stage where 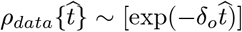 or 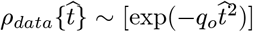, which likely represents a second process, independent of the gradually changing *t_dbl_ doubling time*. An *Enhanced Initial Model (EIM)* is developed here to include this second process. The prior *OFM* methods can then be applied, giving an *Enhanced Orthogonal Function (EOF)* model for this more general case.

### 1.1 Review of Prior Models

The *Initial Model* of Eq. [1.2] is still needed as the first part of the *OFM*. The *Initial Model* starts with measured data end-points {*N_I_*, *N_F_*}, where 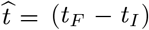 is the largest data time interval so that 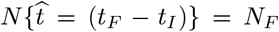. Usually *α_S_* in Eq. [1.2] was chosen first, and *K_o_* or *t_dbl_* adjusted to match the *N_F_* data end-point, using an *Excel^TM^_Goal-Seek* or its equivalent. The final {*K_o_*, *α_S_*} values were the pair with the minimum *root-mean-square* (*rms*) error between the given data and the Eq. [1.2] model.

The above {*K_o_*, *α_S_*} also provides a *t* = 0 estimate for the pandemic start, and gives {*K_A_*, *γ_o_*} as new data fitting parameters:

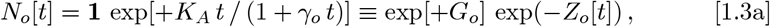

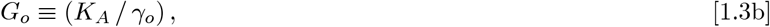

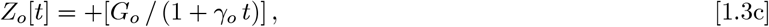

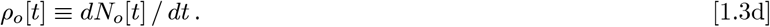

Using (*t_F_* − *t_I_*), with *N_o_*[*t* = *t_I_*] = *N_I_* and *N_o_*[*t* = *t_F_*] = *N_F_*, sets:

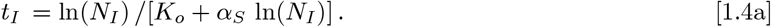

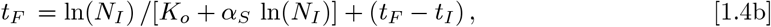

which determines {*K_A_*, *γ_o_*} in *Z_o_*[*t*] for Eq. [1.3c]:

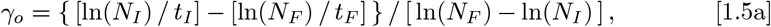

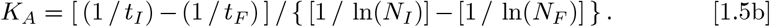

The Eq. [1.4a] value for *t_I_* is what determines the new *t* = 0 point, as an extrapolation for when *N_o_*[*t* = 0] = **1**. In addition:

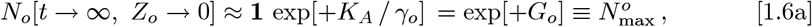

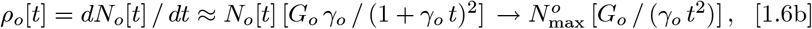

provides an estimate for the total number of cases 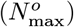 at the pandemic end, and determines a function for the *ρ_o_*[*t*] long-time tail. The {0 < *t* < *t_I_*} period prior to the start of *Social Distancing*, extrapolates what pandemic progress would have looked like, if *Social Distancing* had begun at *t* = 0.

An *Orthogonal Function Model [OFM]* was then developed**^2^** to better model the different observed *ρ_data_*[*t*] shapes, as an improvement of the *Initial Model*:

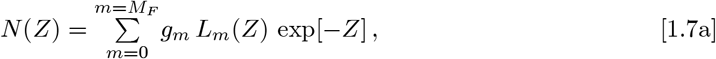

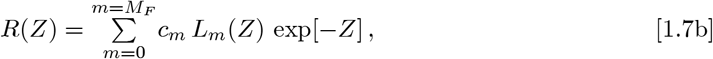

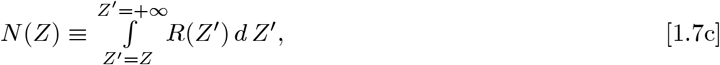

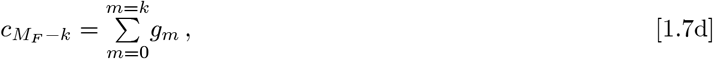

with *L_m_*(*Z*) being the *Laguerre Polynomials*, and *L_m_*(*Z* = 0) = *L_o_*(*Z*) = 1.

Using *Z* = *Z_o_*[*t*] from Eq. [1.3c] gives *N*(*Z*) → *N*(*Z_o_*) and *R*(*Z*) → *R*(*Z_o_*).

The {*g_m_*; *m* = (0, *M_F_*)} constants in Eq. [1.7a] can be arranged in a 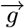 —vector form, with comparable constants for R(Z) from Eq. [1.7b] arranged in a 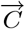 —vector form. For *M_F_* = 2, it allows Eq. [1.7d] to be written as:

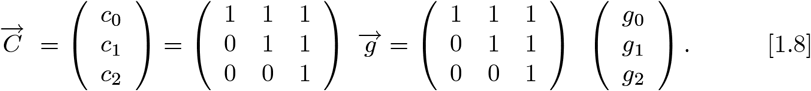

Once these *c_m_* values are determined for *R*(*Z*) in Eqs. [1.7b]-[1.7c], an *OFM* feature is that *c*_0_, by itself, becomes the *OFM* best estimate for the total number of CoVID-19 cases at the pandemic end.

For large enough *M_F_* values, and a monotonic *Z*—function, this *OFM* can provide successively better approximations to almost any given set of *N_data_*[*Z*], with *Z*[*t*] → *Z_o_*[*t*] of Eq. [1.3c] being a specific case.

The *OFM* implicitly uses a *Linear Y-axis*, so its results differ from the *Initial Model* datafit on a *Logarithmic Y-axis*. As an example, compare the *Initial Model* result of *N_o_*(*Z_o_*) = *G*_0_ exp[−*Z_o_*] with the Eq. [1.7a] *OFM* result of *N*(*Z_o_*) = *g*_0_ exp[−*Z_o_*] for *M_F_* = 0. In the *Initial Model, G_o_* is fixed so that *N_o_*(*Z_o_*) exactly matches {*N_I_*, *N_F_*} at the {*t_I_*, *t_F_*} boundaries. In the *OFM*, *g*_0_ = *G_o_* is no longer required, so that the *OFM N*(*Z_o_*) best datafit is not constrained to exactly match {*N_I_*, *N_F_*} at {*t_I_*, *t_F_*}.

The above *R*(*Z*) and *Z*[*t*] gives *N*[*t*] and *ρ*[*t*] as an explicit functions of time:

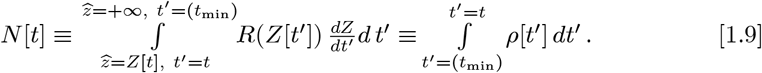

Since Eq. [1.6b] gives *ρ*{*t*} ~ [1/*t*^2^], the aim here is to model faster decaying functions such as *ρ*{*t*} ~ [exp(−*δ_o_t*)] or *ρ*{*t*} ~ [exp(−*q_o_t*^2^)].

### 1.2 Updated *Initial Model* Results for Italy

The *Z_o_*[*t*] model of Eq. [1.3a] was applied to *bing.com* data**^9^** for Italy, starting with 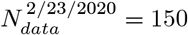 CoVID-19 cases as an early pandemic point, up through June 15, 2020. Here, *t_I_* is when mandatory *Social Distancing* was introduced at 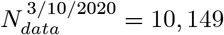; with *t_F_* being when 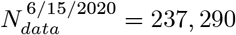. Data prior to *Social Distancing* (*t* < *t_I_*) was excluding from this *Social Distancing* analysis.

**Figure 1** compares the *ρ_data_*[*t*] results with the updated *N_o_*[*t*] and *ρ_o_*[*t*] = *dN_o_*[*t*]/*dt* predictions using *Z_o_*[*t*]. The 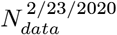 (Day 1) to 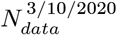 (Day 17) interval was examined for estimating a *t* = 0 pandemic start where *N_data_*[*t* = 0] → **1**. A best fit value of *t_offset_* = 9.10055 days was found, giving *t_I_* = (17 - 9.10055) = 8.89945 days for 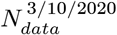 (Day 17), while 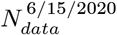 (Day 114) gives *t_F_* = (114 − 9.10055) = 104.89945 days, so that:

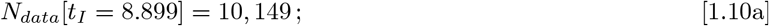

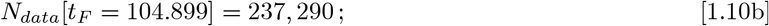

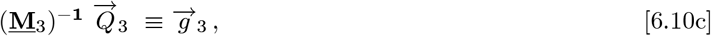

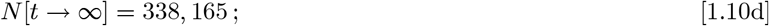

with {*K_A_*, *γ_o_*} ≈ {4.2405, 0.33078} and (*t_F_* − *t_I_*) = 97 days. In **Fig. 1**, the X-axis uses this *t* = 0 point where *N_o_*[*t* = 0] → 1, and it shows what *Social Distancing* effects would have been, if it had been operating throughout the *t* > 0 period. This *ρ_o_*[*t*] prediction still has a much more gradual drop than the data. This discrepancy indicates that a second *Social Distancing* process is operating, besides just the gradual t^ lengthening of the *Initial Model*.

**Figure 1:**
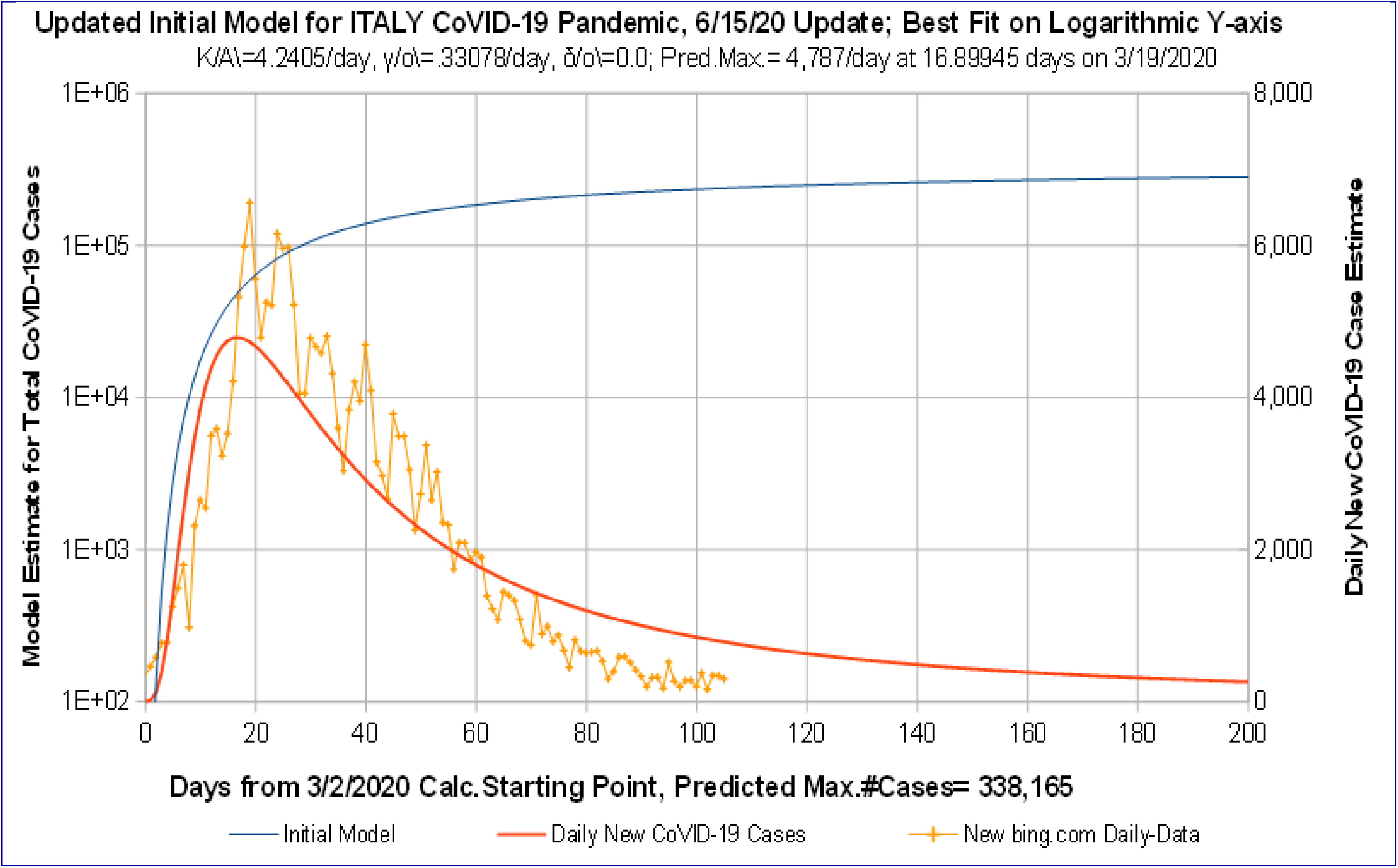
Updated *Initial Model* for ITALY, CoVID-19 data to 6/15/2020. Number of daily CoVID-19 cases calculated as if *Social Distancing* started at 3/2/2020, but only data from 3/10/20 actual *Social Distancing* start, with *N* = 10, 149 cases, was used in calculations.

**Figure 2** compares the *N_o_*[*t*] predictions for this model, to the measured *N_data_*[*t*]. Systematic deviations are evident, with the net *rms* error on a *Logarithmic Y-axis* being *rms_error_* = 0.097828. To cure these defects, an enhanced *Z*[*t*] model is developed next.

**Figure 2:**
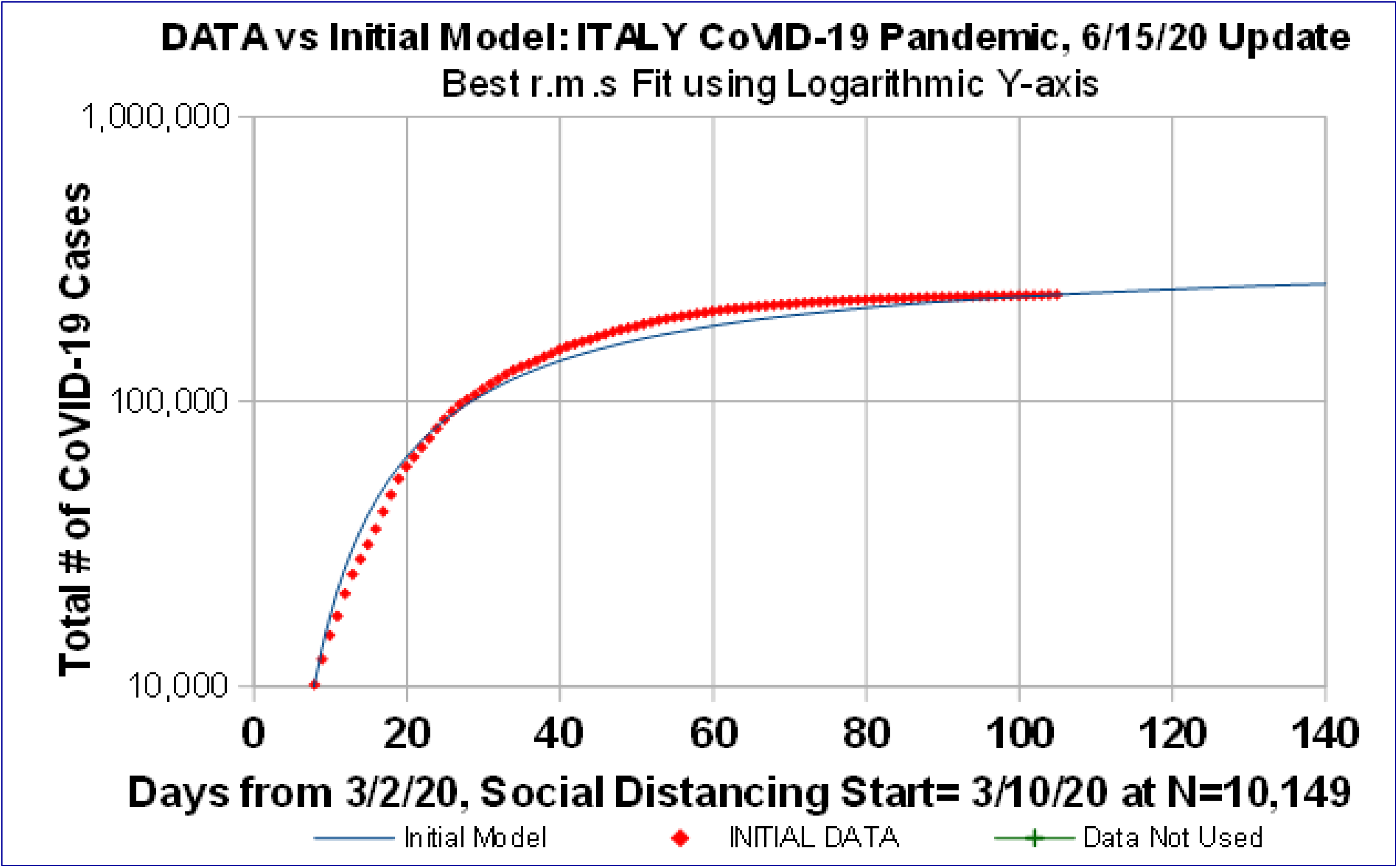
Comparison of *Initial Model* Best-Fit to Measured Data. Best-fit done on *Logarithmic Y-axis*, using data from 3/10/20 actual *Social Distancing* start with *N* = 10, 149 cases, through 6/15/20 with *N* = 237, 290.

## 2 Developing Enhanced *Z*[*t*] Models

To generalize *Z*[*t*] beyond Eq. [1.3c], it is convenient to use the *t* = {0^+^, ∞^−^} domain, and require convergence of *Z*[*t*] → *Z_o_*[*t*] in some limit, along with:

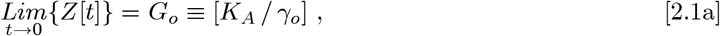

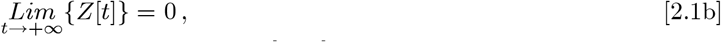

and that the *M_F_* = 0 case of Eq. [1.7a] remains as:

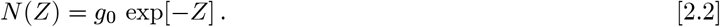

How to choose an appropriate *Z*[*t*], as part of an *Enhanced Initial Model (EIM*), which also allows a *ρ*[*t*] ~ [exp(−*δ_o_t*)] or *ρ*[*t*] ~ [exp(−*q_o_t*^2^)] stage, is examined next. It can be motivated by studying a simple *N_T_*[*t*] test-case, where *ρ_T_*[*t*] itself is a pure exponential decay, as a function of time:

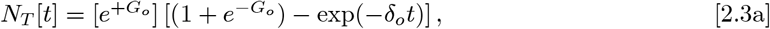

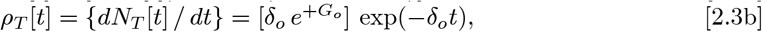

while also preserving *N_T_*{*t* → 0} = 1. Comparing Eq. [2.3a] to the Eq. [2.2], sets *Z_T_*[*t*] for this test-case:

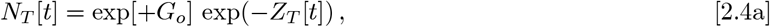

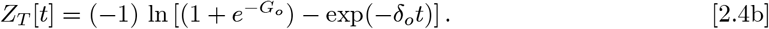

At large times, Eq. [2.4b] gives:

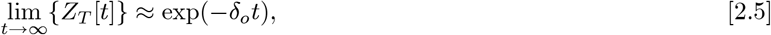

since (− ln[1 − *x*] ≈ *x*) for small x, which shows that if *ρ_T_*[*t*] has an exponential tail, then *Z_T_*[*t*] also has an exponential tail. A simple generalization for *Z*[*t*] in Eq. [1.7a]-[1.7d] would be either:

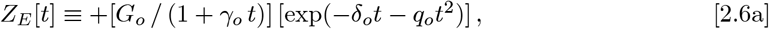

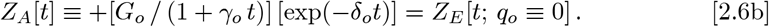

As with Eq. [1.3c], the original CoVID-19 exponential growth factor *K_A_* remains only as part of the *G_o_* scaling factor, while the {*δ_o_* → 0} limit of Eq. [2.6b] converges back to the Eq. [1.3c] *Initial Model*.

Using Eq. [2.6a] for *Z_E_*[*t*] in the Eq. [2.2] *N*(*Z_E_*) example gives:

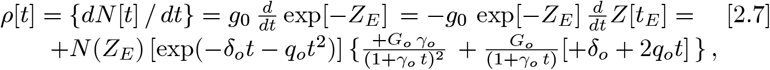

which exhibits the following variety of long-time limits:

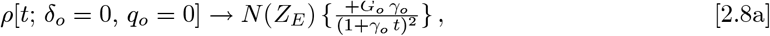

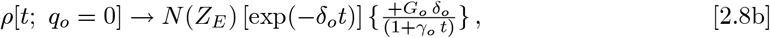

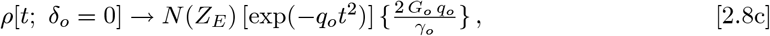

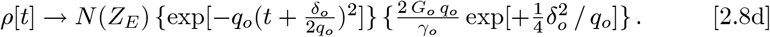

Here, any *q_o_* ≠ 0 Gaussian component in *Z_E_*[*t*] gives a *ρ*[*t*] tail that is also a pure Gaussian. An exponential component (*q_o_* ≡ 0) in *Z_E_*[*t*] gives a time- modified exponential *ρ*[*t*] tail, while having {*q* ≡ 0, *δ_o_* ≡ 0} in *Z_E_*[*t*] gives the prior *ρ*[*t*]^~^(1/*t*^2^) result of Eq. [1.6b].

## 3 Pandemic Fast vs Slow Shutoffs

Each *Z_E_*[*t*] function modifies *N*[*t*] predictions for the pandemic start, pandemic end, and the mid-range where *ρ*[*t*] has its pandemic peak. The Eq. [2.6a] *Z_E_*[*t*] function especially alters the calculated CoVID-19 pandemic tail. For either *q_o_* ≠ 0 or *δ_o_* ≠ 0, Eqs. [2.6a]-[2.6b] gives a pandemic *Fast Shutoff*, compared to the gradually decreasing *Z_o_*[*t*] of Eq. [1.3c] in the *Initial Model*, which is a pandemic *Slow Shutoff*. However *Z_o_*[*t*] from the *Initial Model*, and *Z_A_*[*t*] from Eq. [2.6b] both gave long-term *ρ*[*t*] tails that decay much slower than the *q_o_* ≠ 0 Eq. [2.6a] Gaussian.

If data does not show evidence of a Gaussian pandemic *Fast Shutoff*, assuming the post-peak *ρ*[*t*] data will be Gaussian is likely to provide optimistically inaccurate *N*[*t*] predictions for CoVID-19 pandemic evolution. Apparently, this is exactly what was done by the *University of Washington IHME (Institute of Health Metrics and Evaluation)* in their widely publicized initial preprint**^3^** of 27 March 2020, with this Gaussian model continuing throughout their subsequent updates**^4-6^** up through 29 April 2020.

*IHME* changed everything in their 4 May 2020**^7–8^** update. They no longer used *ρ*[*t*] Gaussian tails, and it doubled or tripled their predicted CoVID-19 pandemic death rates. Thus, unless the post-peak *ρ*[*t*] exhibits Gaussian behavior, the *Z_A_*[*t*] with *δ_o_* =0 is likely the most important modification to *Z_o_* [t], which is the pandemic *Fast Shutoff* model used here.

Using *Z_A_*[*t*], the pandemic *Fast Shutoff* can be extrapolated to calculate a pandemic start point where *N*[*t* = 0] = 1. We then examine if this *Z_A_*[*t*] must also carry over to the *ρ*[*t*] long-term tail,

Since the long-term low *p_data_*[*t*] tail may differ among localities, and is not well known, the *δ_o_* ≠ 0 case of Eq. [2.6b] could end with a *Slow Shutoff*, giving:

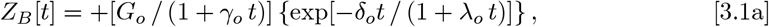

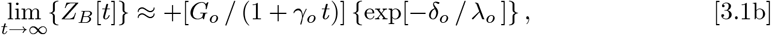

with *G_o_* as in Eq. [2.1a]. This Eq. [3.1a] *Z_B_*[*t*] function has the {*δ_o_*, *λ_o_*} *Mitigation Measure* operating at the start of *Social Distancing*, but reverting to the *Initial Model* in the long-time limit. Combining Eq. [2.6b] and Eq. [3.1a] cases gives this *Enhanced Initial Model (EIM)* equation:

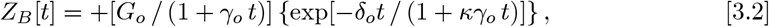

where *κ* = 0 is a pure exponential, and *κ* = 1 has a modified tail that includes its own long-term shutoff. Comparing *κ* = {0,1} in Eq. [3.2] provides a simple test for which model matches CoVID-19 data better in any locality. Any other *κ* > 0 value then recovers the more general A_o_ = *κγ_o_* case. This Eq. [3.2] *Z_B_*[*t*] replaces *Z_o_*[*t*] of Eq. [1.3c], and its *EIM* companion *N_B_*[*t*] is:

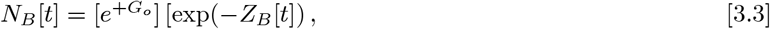

while using *Z*[*t*] → *Z_B_*[*t*] in Eqs. [1.7a]-[1.7d] gives an *Enhanced Orthogonal Function [EOF]* model.

## 4 Finding {*K_A_*, *γ_o_*, *δ_o_*} for *Z_B_*[*t*] from Data

If *δ_o_* = 0, the prior Eqs. [1.4a]-[1.5b] for {*K_A_*, *γ_o_*, *δ_o_* = 0} and {*t_I_*, *t_F_*} can be used, with the initial best-fit {*K_o_*, *α_S_* } values determined by minimizing the *rms* error between Eq. [1.2] and the measured data on a *Logarithmic Y-axis*. Unfortunately, Eqs. [1.4a]-[1.5b] cannot be used when *δ_o_* = 0, although finding a good {*K_A_*, *γ_o_*, *δ_o_*} starting point is still needed for the *EIM*:

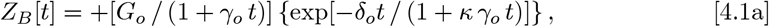

prior to any *EOF* analysis. The *κ* = 1 case also has this special symmetry:

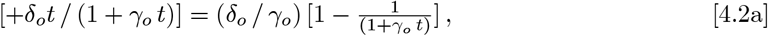

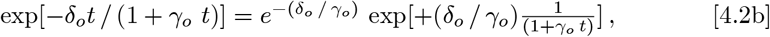

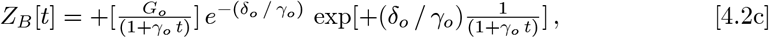

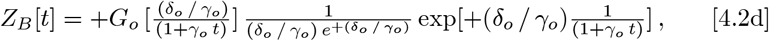

which can re-written as:

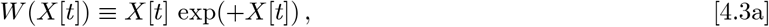

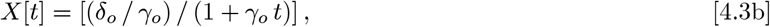

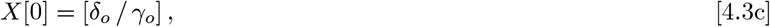

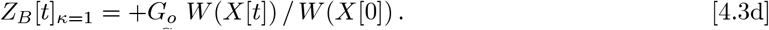

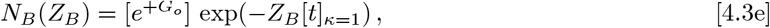

while the *κ* = 0 case is:

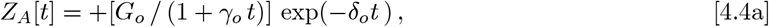

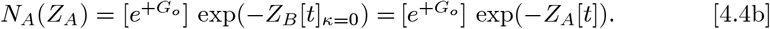

For *κ* = {0, 1}, the *t* = 0 point, *G_o_* from Eq. [2.1a], and the *N_I_*(*t_I_*) and *N_F_*(*t_F_*) initial and final points, give these equations to help set {*K_A_*, *γ_o_*, *δ_o_*}:

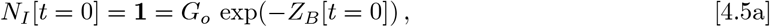

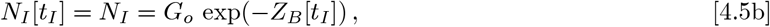

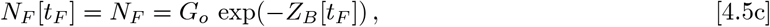

for the *EIM*. Minimizing the *rms* error between the Eq. [3.3] {*K_A_*, *γ_o_*, *δ_o_*, *t_I_*} functions and measured data on a *Logarithmic Y-axis* can be done as follows. Start with estimated values for 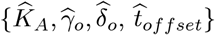 in:

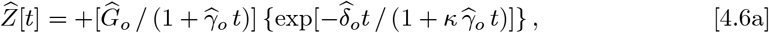

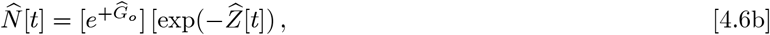

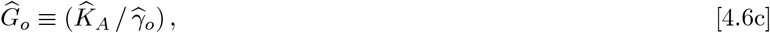

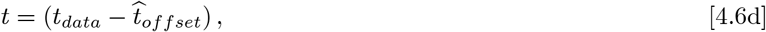

where *t_data_* is the data start time. Set a preliminary value for 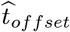 first, to fix the time scale for the *N_data_* [t] measured values:

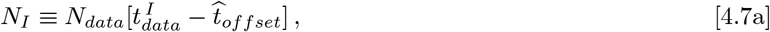

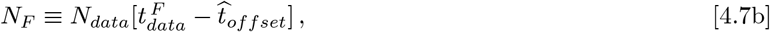

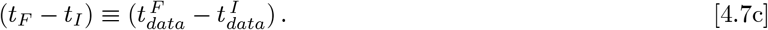

Next, pick values for 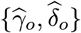 for 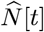 in Eq. [4.6b], allowing direct comparison between *N*[*t*] and 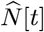 at each data point:

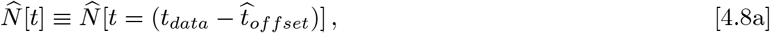

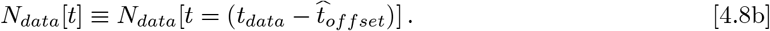

The resulting calculated values for both 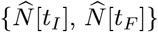 can often be much too high or low, compared to the {*N_I_*, *N_F_*} measured data, but those values can be renormalized to:

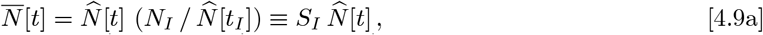

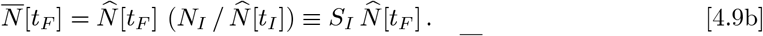

Here, *S_I_* is the renormalization coefficient, and 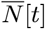 allows easy comparison to the measured *N_data_*[*t*] since 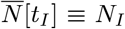. Given 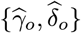, the 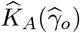 value that is needed to obey 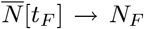 can be set by using *Excel*™*_Goal-Seek* or its equivalent, which also sets a particular *S_I_* value. Next, the 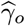 value is adjusted to find the specific {*K_A_*, *γ_o_*} parameter pair that gives *S_I_* = 1. This process is needed because these 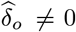 cases do not allow easy determination of *t_I_* as in Eq. [1.4a], or for {*K_A_*, *γ_o_*}, given *t_I_*, as in Eqs. [1.5a]-[1.5b].

The *rms* error on a *Logarithmic Y-axis*, between this 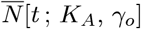 and the *N_data_*[*t*] is one of many 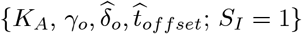 choices. The minimum rms error among all these *S_I_* = 1 cases and the *N_data_*[*t*], when varying 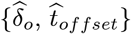 gives the best {*K_A_*, *γ_o_*, *δ_o_*, *t_offset_*} values for Eqs. [4.3a]-[4.3e].

## 5 *Enhanced Initial Model [EIM]* Results for Italy

The Eq.[1.3a] *Initial Model* results were shown in **Figs. 1-2**. Mandatory *Social Distancing* was introduced at 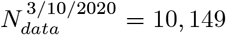 which is the *t_I_* data point, with 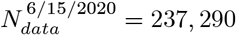 being the *t_F_* data point. The *EIM* was then applied to the same data, to highlight the improvements that can be obtained from using the *EIM* of *Z_A_*[*t*] and *N_A_*(*Z_A_*), in place of *Z_o_*[*t*] and *N_o_*(*Z_o_*).

**Figures 3-4** show the resulting *EIM* best-fits for *Z_A_*[*t*] and *N_A_*(*Z_A_*), along with using a *ρ_A_*[*t*] tail that is a pure exponential decay.

**Figure 3:**
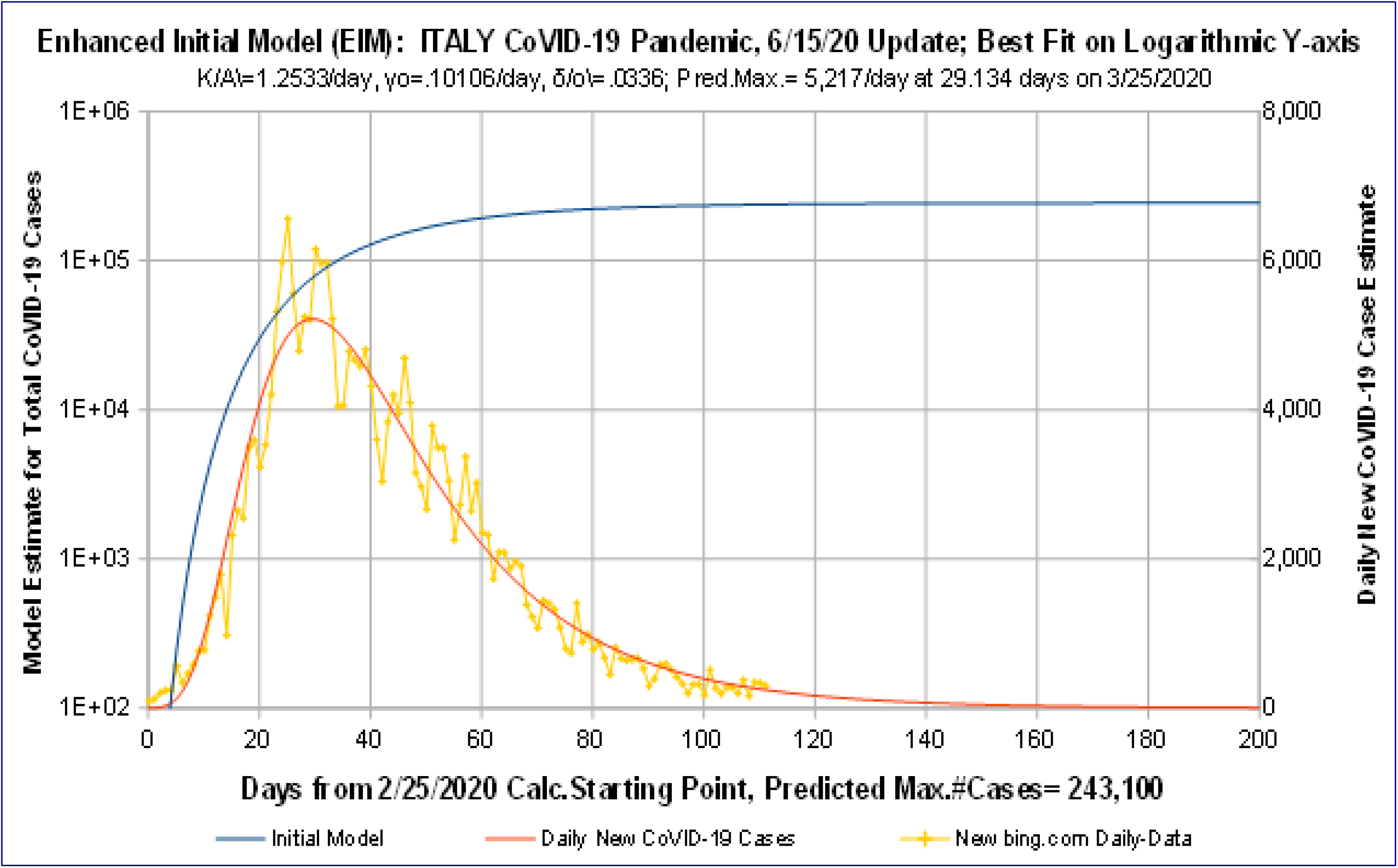
*Enhanced Initial Model (EIM)* for ITALY CoVID-19 data to 6/15/20. *EIM* best fit with *N_A_*[*t*] ~ exp(−*Z_A_*[*t*]) using enhanced *Z_A_*[*t*] function having an exponential decay. Adding in exponential decay term gives significantly improved fit, compared to prior *OFM*.

**Figure 4:**
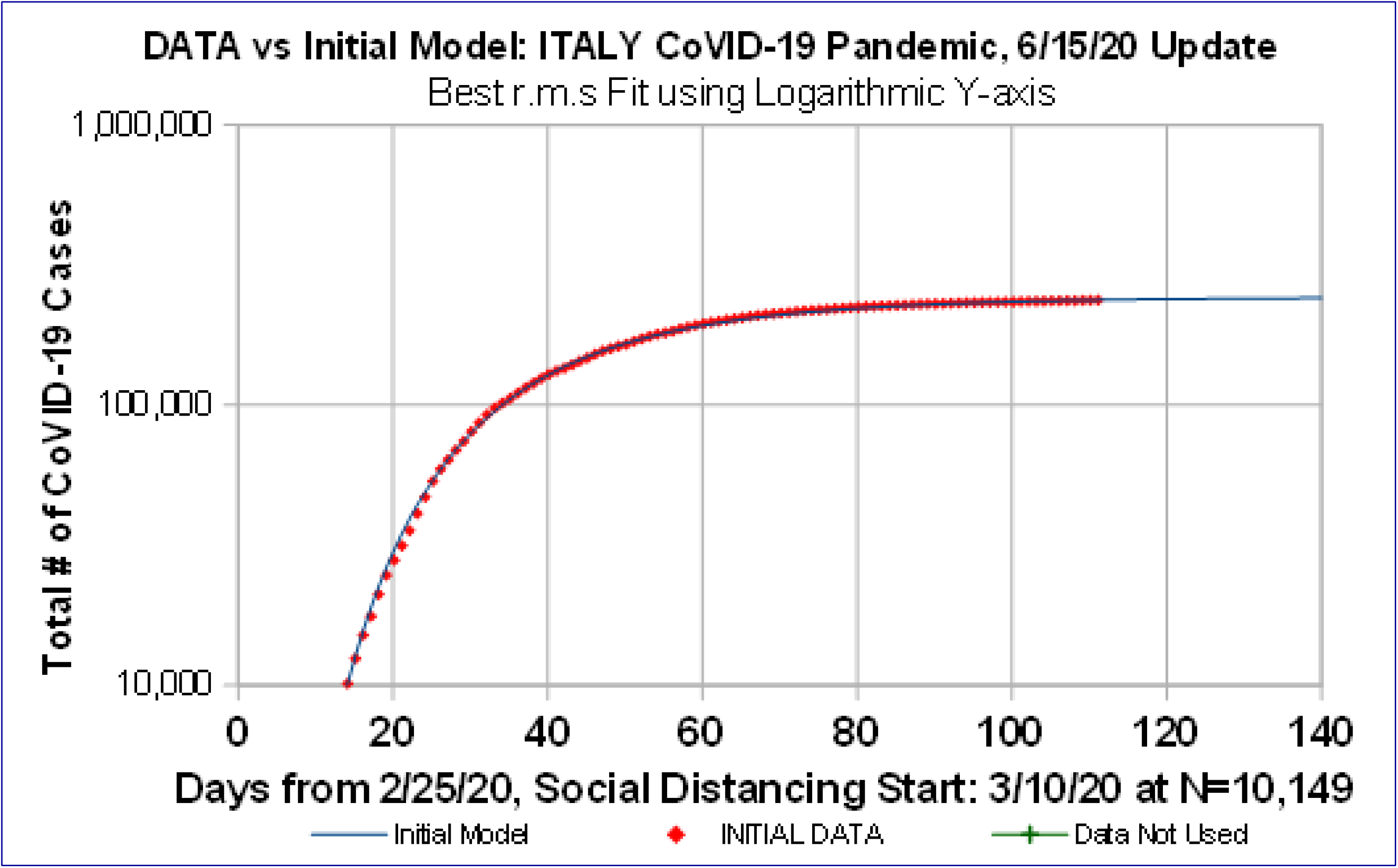
*Enhanced Initial Model (EIM)* datafit for ITALY CoVID-19 data to 6/15/20. *EIM* best fit with *N_A_*[*t*] ~ exp(−*Z_A_*[*t*]) using enhanced *Z_A_*[*t*] function having an exponential decay. Datafit minimizing *rms* error on *Logarithmic Y-axis* gives significant improvement vs *OFM*.

For the *EIM*, a new best estimate of *t_offset_* = 2.866 days was found within the 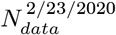 (Day 1) to 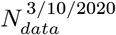 (Day 17) data, setting the *EIM* t = 0 point. Then *t_I_* = (17 — 2.866) = 14.134 days, while 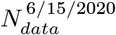 (Day 114) gives *t_F_* = (114 — 2.866) = 111.134 days, along with:

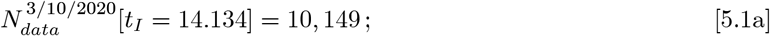

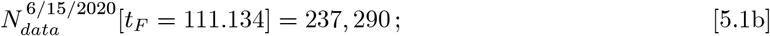

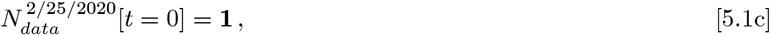

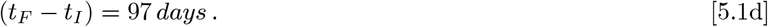

This X-axis *t* = 0 point is a hypothetical *EIM* pandemic starting point, if *Social Distancing* had been operating throughout the initial CoVID-19 period. **Figure 3** has a predicted CoVID-19 pandemic peak of ^~^5, 217 / day at *t* = 29.134 days on 3/25/2020, with ^~^243,100 total cases at the pandemic end. This datafit has > 4*X* error reduction over the *δ_o_* = 0 case, as summarized next:

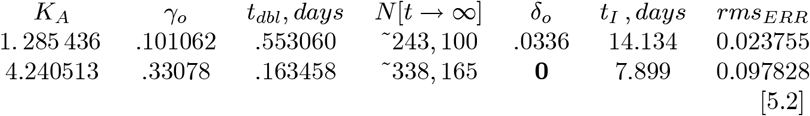

The *EIM* with *Z_A_*[*t*] and *δ_o_* = 0.0336 gives a *ρ_A_*[*t*] curve that is in excellent agreement with the **Fig. 3** measured *ρ_data_*[*t*] data. Comparing *N_A_*[*t*] and *N_data_*[*t*] in **Fig. 4** also shows an excellent match over the whole *Logarithmic Y-axis* range, used in the *rms* error minimization.

The **Figs. 3-4** *κ* = 0 results were then compared to the *κ* =1 case, using Zb[t] and Nb(Zb) of Eqs. [3.2]-[3.3]. The *κ* = 1 case has a *Social Distancing* factor that gradually turns off the *EIM* exponential decay. The resulting *rms* error best fits converged to *δ_o_* → 0, as follows:

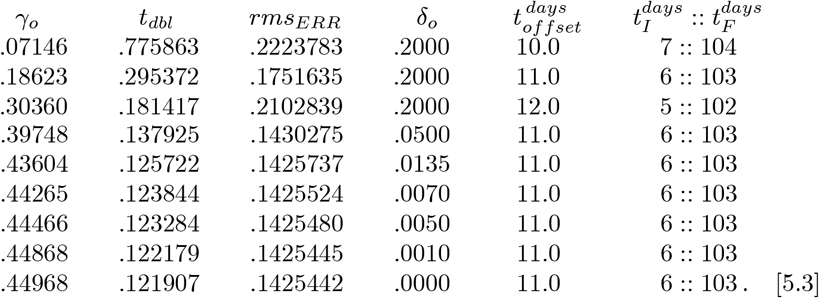

This *κ* = 1 result shows that the process giving rise to exponential tails in the *ρ_data_*[*t*] for a pandemic *Fast Shutoff* is independent of the *Social Distancing* process that gradually lengthens the *t_dbl_ doubling time*, as measured by *γ_o_*. Comparing the *κ* = {0,1} cases shows that the pure exponential tail with *Z_A_*[*t*] = *Z_B_*[*t*]*_κ_*_=0_ matches the *ρ_data_*[*t*] data best.

## 6 *Enhanced Orthogonal Functions* for Italy

Any monotonic *Z*[*t*] can convert measured *N_data_*[*t*] data into *N_data_*(*Z*). Using the Eq. [2.6b] *Z_A_*[*t*] → *Z*[*t*] in Eqs. [1.7a]-[1.7d] extends the *EIM* into an *EOF* model, where Eq. [1.9] gives:

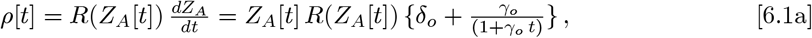

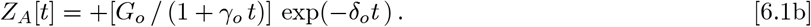

The Eq. [5.2] *δ_o_* ≠ 0 entries and Eq. [5.1a]-[5.1d] boundary conditions give:

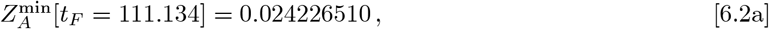

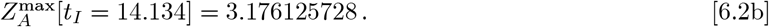

These data-driven *Z_A_*[*t*] limits are used next, along with *Z*[*t*] → *Z_A_*[*t*] in:

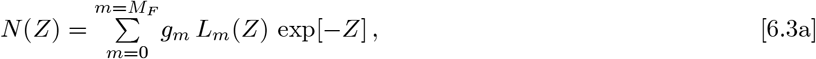

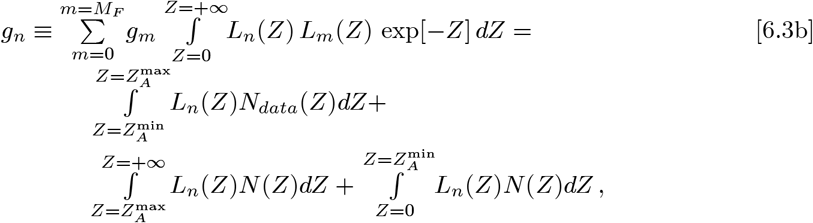

where *m* = {0, *M_F_*} sets how many terms are in the Eq. [6.3a] series.

Generally *M_F_* = 2 is used here. The *L_m_*(*Z*) are the *Laguerre Polynomials*, with the first few *L_m_*(*Z*) being:

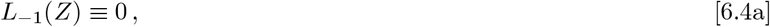

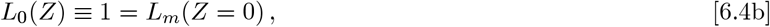

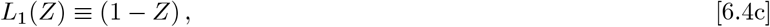

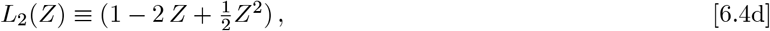

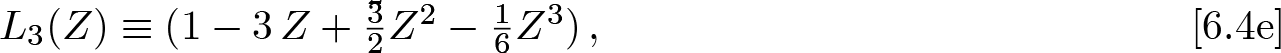

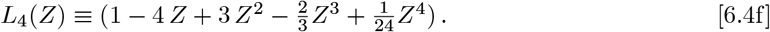

Some important properties of the *Laguerre Polynomials* are:

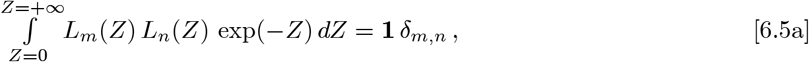

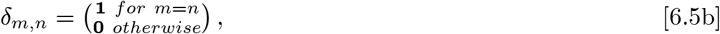

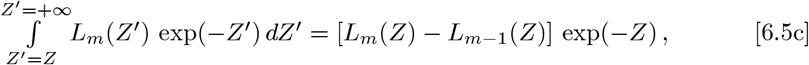

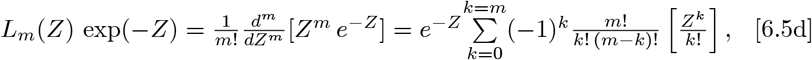

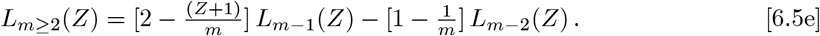

Here Eq. [6.5a] defines an *orthogonal function set*. The “**n**!” (**n***-factorial*) in Eq. [6.5d], for **n** an integer, is defined as the product:

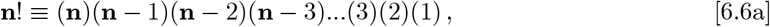

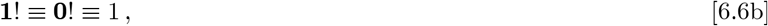

where *factorials* with negative integers are not allowed. For *M_F_* > 2, the following equations developed by Watson**^10^**, and improved by Gillis and Weiss**^11^**, helps in evaluating Eq. [6.3b]:

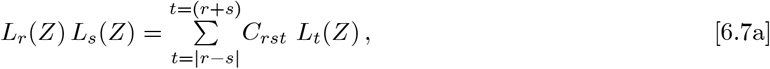

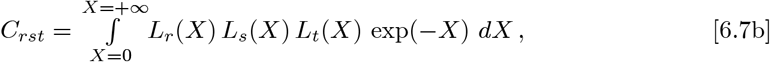

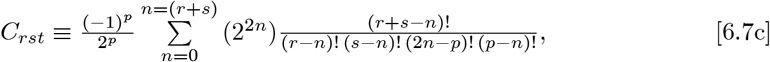

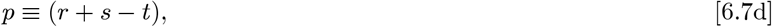

where ALL terms in the Eq. [6.7c] sum for *n* = {0, (*r* + *s*)} have an implicit requirement that all negative *factorial* arguments are excluded**^10–11^**.

Since the *N*(*Z*) of Eq. [6.3a] has {*g_m_*; *m* = (0, *M_F_*)}, and *N*(*Z*) also appears in each *g_n_*-equation of Eq. [6.3b], how to determine each *g_m_* by itself, can be done as follows. First define:

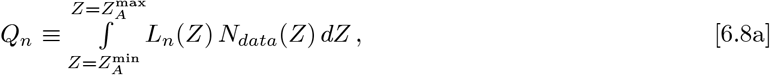

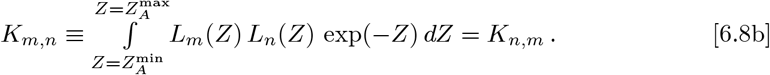

When the *N_data_*(*Z*) is comprised of *j* = {1, 2, …*J*} discrete values between 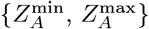 with each *Z_j_* having an 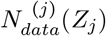 value, the Eq. [6.8a] integral needs to be replaced by a sum. Let *Z*_0_ = *Z*_1_ and *Z*_J+1_ = *Z_j_*, the *Q_n_* replacement for Eq. [6.8a] is then:

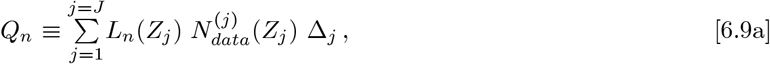

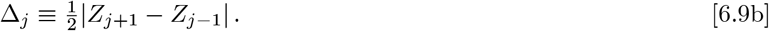

Eq. [6.3b] can then be re-written as a 3 × 3 matrix **M**_3_, which relates a data-driven 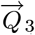-vector to a resultant 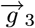-vector:

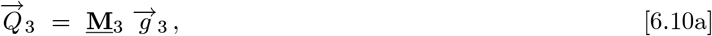

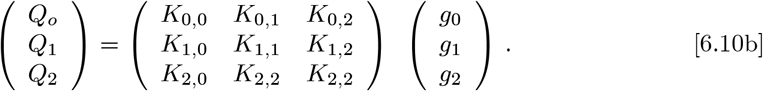

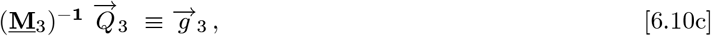

where (**M**_3_)^−1^ is the matrix inverse of **M**_3_. When 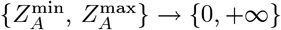, this **M**_3_ becomes the Identity Matrix. The following *K_m.n_*(*Z*) integrals set *K_m,n_*:

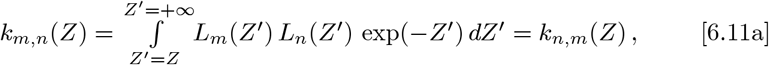

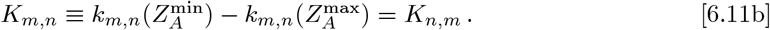

The *K_m.n_*(*Z*) integrals can be determined using Eq. [6.5c], which gives:

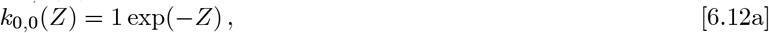

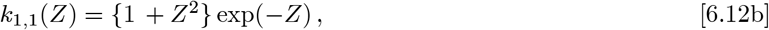

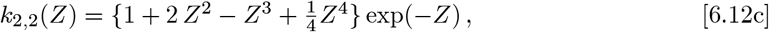

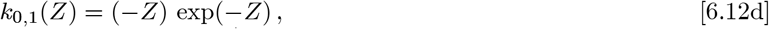

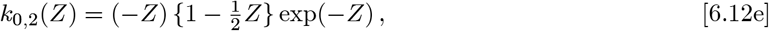

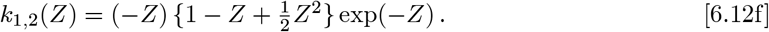

The extract {*g*_0_, *g*_1_, *g*_2_}, the 3 × 3 symmetric **M**_3_ matrix needs inversion:

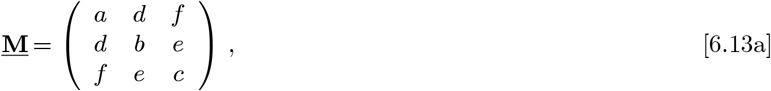

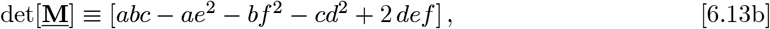

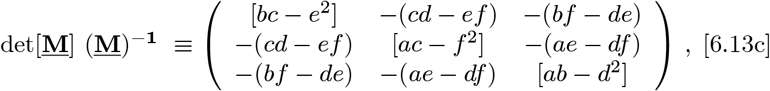

which determines {*g*_0_, *g*_1_, *g*_2_} from the {*Q*_0_, *Q*_1_, *Q*_2_} data. A best-fit *N*(*Z*) for *Z* = {0^+^,∞^−^} results, along with an equivalent fit for *R*(*Z*) using Eq. [1.7d].

Instead of having to find the best {*g*_0_, *g*_1_, *g*_2_} triplet, one could find the best 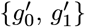 by just using using {*Q*_0_, *Q*_1_} and an **M**_2_ sub-matrix; or one could find the best 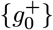 by itself by just using {*Q*_0_} and an **M**_1_ sub-matrix:

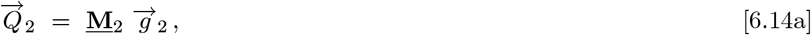

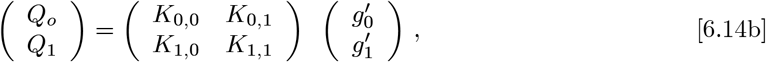

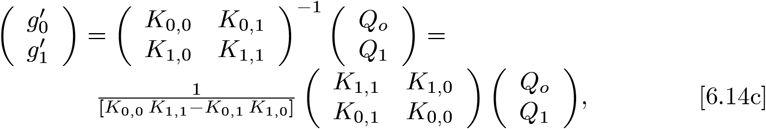

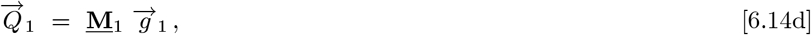

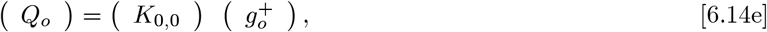

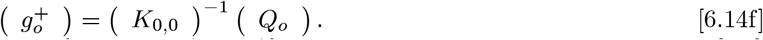

Once the {*g_m_*; *m* = (0, *M_F_*)} constants are found and used in Eq. [1.8], its *c*_0_ value provides the new *EOF* estimate for the predicted total number of CoVID-19 cases at the pandemic end, refining the initial Eq. [5.2] *N*[*t* → ∞] *EIM* value.

## 7 *EOF* Model Results for Italy

The *EOF* model starts with the *EIM* of Eq. [2.6b] using *Z_A_*[*t*], and the *bing.com* Italy data**^9^**, which gives:

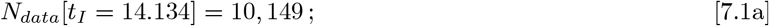

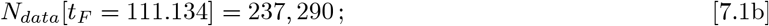

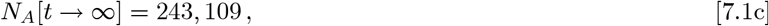

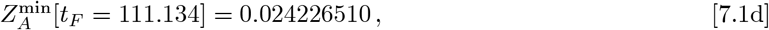

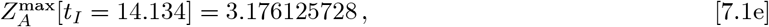

via Eqs. [5.1a]-[5.1d], [5.2], and [6.2a]-[6.2b]. For these 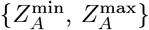 values, with (*t_F_* − *t_I_*) = 97 days, the **M**_3_ matrix of *K_m,n_* entries, via Eq. [6.8b], is:

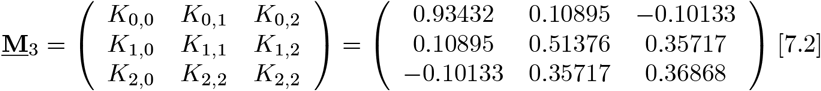

It has a rather small det[**M**_3_] = 0.04024218 value, with an inverse of:

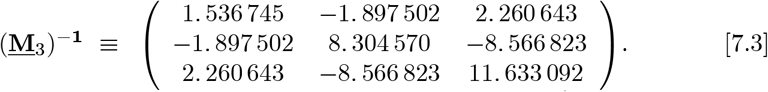

A convolution of *L_m_*(*Z_A_*) functions with the measured 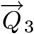 dataset vector of Eqs. [6.9a]-[6.9b], along with the above (**M**_3_)^−1^, gives this final 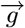 –vector^12^:

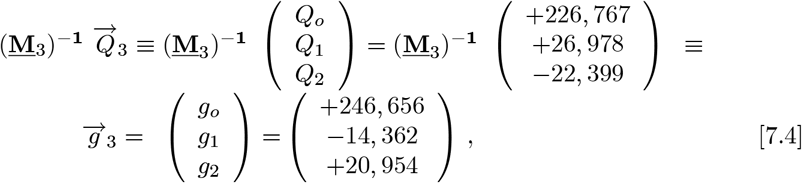

determining the constants for *N*(*Z_A_*) in Eq. [1.7a]. The coefficients for *R*(*Z_A_*), which sets the predicted number of daily new CoVID-19 cases, are:

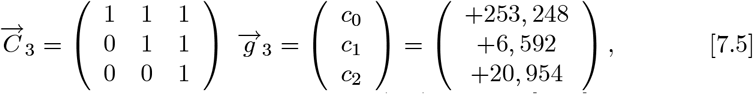

determining the constants needed for *R*(*Z_A_*) in Eq. [1.7b]. Using these {*g*_0_, *g*_1_, *g*_2_} values along with Eq. [1.8] gives:

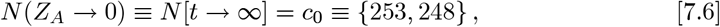

as a new predicted total number of CoVID-19 cases at the pandemic end for the *EOF* model, which is a ^~^4.17% or 10, 139 increase in the number of cases, compared to the *EIM* value of Eq. [7.1c].

Using Eq. [6.1b] for *Z_A_*[*t*], and substituting the Eq. [7.3] 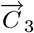 values into Eq. [1.7b] gives *R*(*Z_A_*). The *ρ*[*t*] in Eq. [6.1a] is derived from *R*(*Z_A_*) using Eq. [1.7b], with the resulting *EOF ρ*[*t*] plotted in **Figure 5**, along with the *t* > *t_I_* raw data for the daily new CoVID-19 cases.

**Figure 5:**
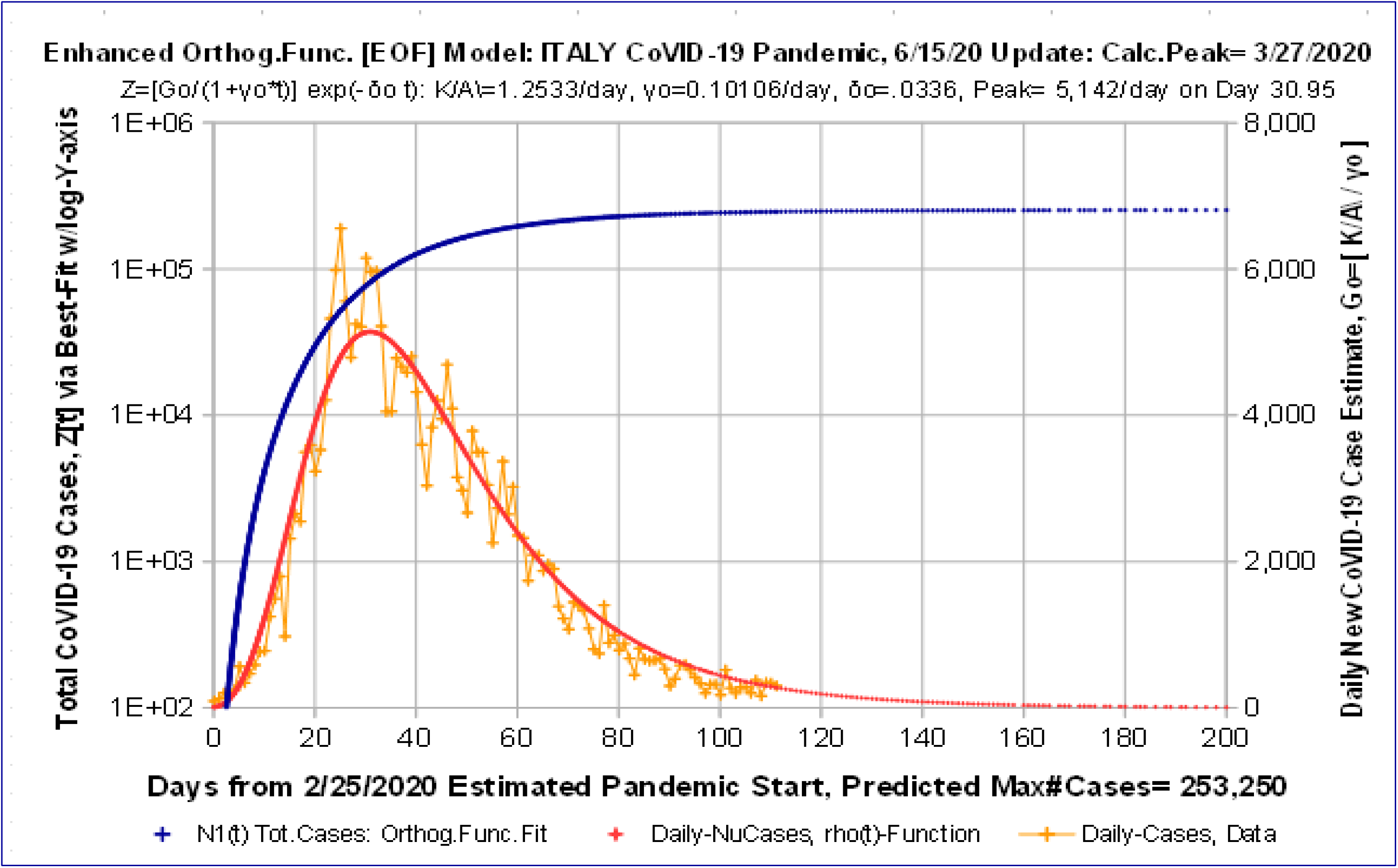
*Enhanced Orthogonal Functions (EOF)* for ITALY CoVID-19 data to 6/15/20. Orthogonal functions give additional parameters for further datafit improvement, using a 3-term series (*M_F_* = 2). Result shows *EIM*, by itself, provides most of the improvement.

The **Figure 5** *EOF* model also gives a *t* < *t_I_* extrapolation, which shows what the combination of processes would look like, if they all had been operating continuously from the CoVID-19 pandemic start. The companion *N*[*t*] analytic result, along with the *t* > *t_I_* raw data for the total number of CoVID-19 cases is show in **Figure 6**.

**Figure 6:**
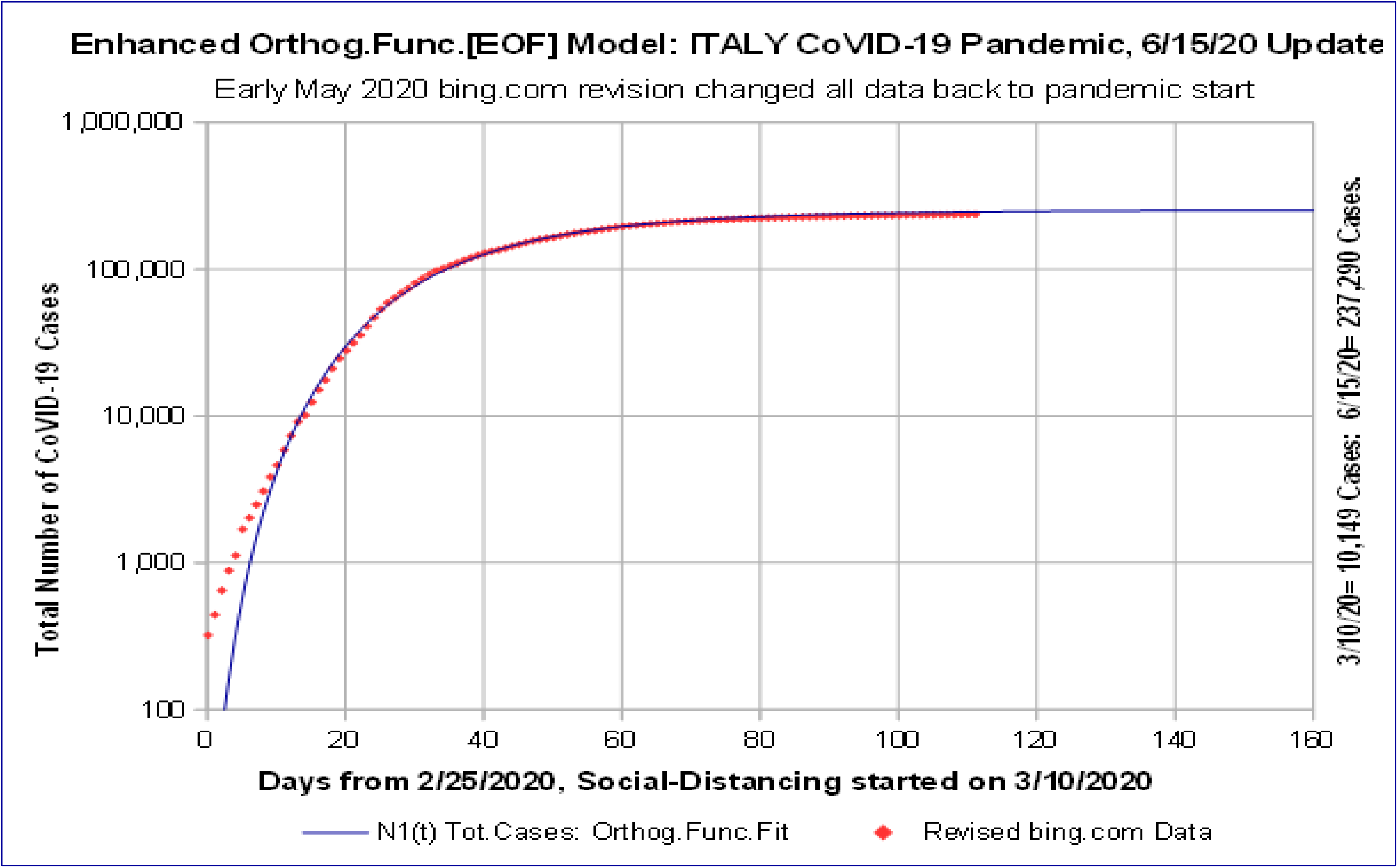
*Enhanced Orthogonal Functions (EOF)* for ITALY CoVID-19 data to 6/15/20. Data for total number of CoVID-19 cases versus time, compared with 3-term *EOF* model for *N*(*Z_A_*[*t*]) shows excellent match after *Social Distancing* start at *N* = 10, 149.

Comparing the size and timing of the *ρ*[*t*] pandemic peak, and its Day 200 value, between the *EIM* (**Figs. 3-4**) and *EOF* model (**Figs. 5-6**), gives:

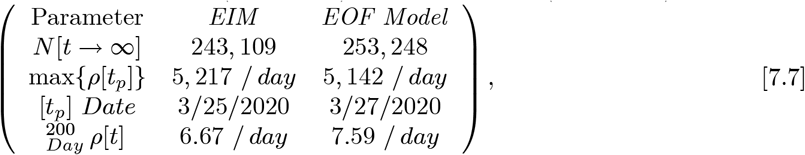

showing the *EOF* model predicts more cases total and more daily new CoVID-19 cases at Day 200, as well as modifying the pandemic peak predictions.

While the above analysis used *M_F_* = 2, with the Eq. [7.4] 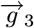 setting the best {*g*_0_, *g*_1_, *g*_2_} values, this *EOF* model also provides estimates for the simpler *M_F_* = {0, 1} cases, as outlined by Eqs. [6.14a]-[6.14f]. For *M_F_* = 1, the best two 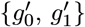 values were gotten by only using {*Q*_0_, *Q*_1_} and an **M**_2_ sub-matrix of **M**_3_. For *M_F_* = 0, the best 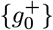 by itself is derived by using {*Q*_0_} and the **M**_1_ sub-matrix. These alternative estimates give:

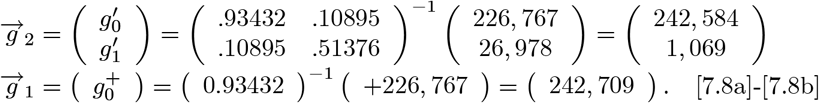

These additional calculations give the following progression of estimates for *N*[*t* → ∞], which is the final number of CoVID-19 cases at the pandemic end:

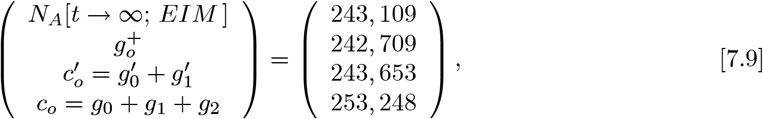

based on increasing the number of data fitting parameters used with the original data. This summary shows the *N*[*t* → ∞] projections are fairly stable, with an average and 1σ standard deviation:

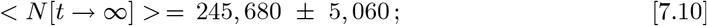

among these different calculations, where 1σ is ^~^2.06% of the overall average.

Comparing the results among **Figs. 1-6** also highlights these items:

a. All *ρ*[*t*] functions have a sharp rise, and slower decreasing tail. The fastest changing *ρ_data_*[*t*] tail, as in the Italy CoVID-19 *(Fast Shutoff*) data, was successfully modeled by adding in an exponential term, as in Eq. [6.1b].
b. The datafits in **Fig. 4** and **Fig. 6** show that the extra parameters in the *EIM* and *EOF* model fits the *ρ_data_*[*t*] shape progressively better.
c. The *EOF* model shows only relatively small changes of ^~^2.06% in the *N*[*t* → ∞] limits (Eq. [7.10]), as an estimate of uncertainty in the *EIM*.
d. The *Enhanced Initial Model (EIM)* function captures much of the progression to a pandemic *Fast Shutoff*, as seen in the Italy data.

The *ρ*[*t*] tail may still differ from these predictions, due to factors such as:

i. The CoVID-19 dynamics may change in the long-term low *ρ*[*t*] regime;
ii. A “second wave” or multiple waves of *ρ*[*t*] resurgence may occur, which are beyond the scope of this CoVID-19 pandemic modeling.

## 8 Summary and Conclusions

The early stages of the CoVID-19 coronavirus pandemic starts off with a nearly exponential rise in the number of infections with time. Defining *N*[*t*] as the expected total number of CoVID-19 cases vs time, this basic function:

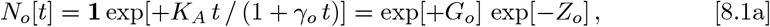

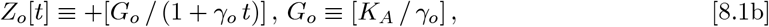

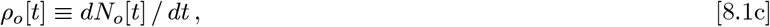

models *Social Distancing* effects as a gradual lengthening of the pandemic growth *doubling time*, which enables pandemic shutoff with only a small population of infected persons. The Eq. [8.1b] *Z_o_*[*t*] was our *Initial Model***^1^**, and gives a CoVID-19 *Slow Shutoff* with a long-term *ρ_o_*[*t*]~[1/t^2^] tail. Previously we showed**^1–2^** that this *Z_o_*[*t*] model fits many *N_data_*[*t*] and *ρ_data_*[*t*] cases.

However, some data had a CoVID-19 *Fast Shutoff*, with a *ρ_data_*[*t*] ~ [exp(−*δ_o_t*)] exponential tail, such as in Italy**^9^**, where a Gaussian tail *ρ*[*t*] ~ [exp(−*q_o_t*^2^)] would have decreased too quickly. An *Enhanced Initial Model (EIM)* was developed here, using this *Z_A_*[*t*] function:

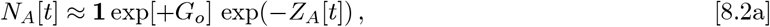

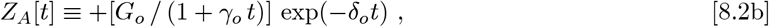

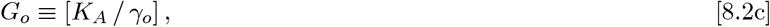

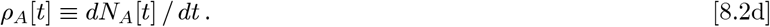

We also examined if the exp(−*δ_o_t*) exponential decay could also be subject to a *Slow Shutoff*, giving exp[−*δ_o_t* / (1 + *γ_o_t*)] instead of exp(−*δ_o_t*), but that did not match the Italy data. To allow more data fitting parameters beyond just {*K_A_*, *γ_o_*, *δ_o_*}, an orthogonal function method was developed**^2^**:

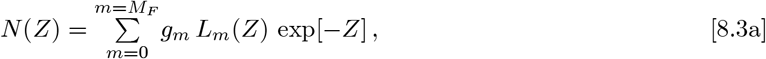

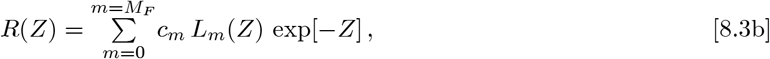

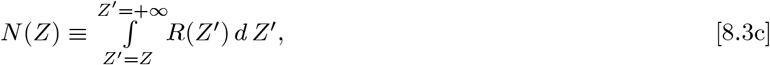

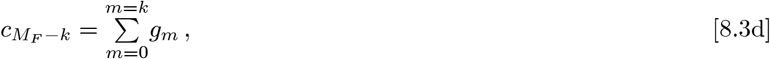

which is applicable to a generic *Z*[*t*] function, with *N*[*t*] = *N*(*Z*[*t*]), where *Z*[*t*] → *Z_o_*[*t*] and *Z*[*t*] → *Z_A_*[*t*] are special cases. Larger *M_F_* with more {*L_m_*(*Z*); *m* = (0, + *M_F_*)} terms can match almost any *arbitrary function*, enabling fits to a variety of *N*[*t*] and *ρ*[*t*] shapes. The {*g_m_*; *m* = (0, + *M_F_*)} are constants determined from each dataset. The *L_m_*(*Z*) are the *Laguerre Polynomials*, with several important properties given in Eqs. [6.4a]-[6.5e].

Using *Z_A_*[*t*] in Eqs. [8.3a]-[8.3d] results in this *Enhanced Orthogonal Function [EOF]* model, which is applicable to both *Slow* or *Fast Shutoff* CoVID-19 pandemic data. The *ρ*[*t*] expected number of daily new CoVID-19 cases is:

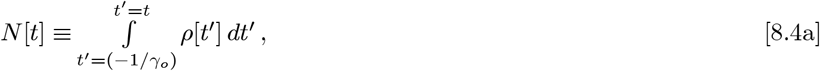

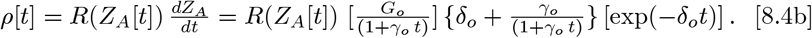

Methods were developed to derive the {*K_A_*, *γ_o_*, *δ_o_*} values, and to determine the {*g_m_*; *m* = (0, + *M_F_*)} and {*c_m_*; *m* = (0, + *M_F_*)} constants from data. Whereas our *Initial Model* and *EIM* were *M_F_* = 0 cases, the *M_F_* = 2 case was used here to examine the Italy CoVID-19 data, as an *EOF* model example.

The *bing.com* data for Italy up to ^~^6/15/2020 was then analyzed, with **Figures 3-6** giving the new Italy results. Both the *EIM* and the *EOF* model provided good datafits, giving similar *N*[*t* → ∞] results for the final number of CoVID-19 pandemic cases, differing by only ^~^2% at the 1σ level.

The *ρ*[*t*] post-peak behavior best indicates if a *δ_o_* ≠ 0 model (CoVID-19 pandemic *Fast Shutoff*) is applicable. The *δ_o_* ≠ 0 case likely is a second *Social Distancing* process, that operates along with, but is independent of the gradual pandemic *doubling time* changes. That *doubling time* change gives rise to a CoVID-19 pandemic *Slow Shutoff* (*γ_o_* ≠ 0), and that process still operates concurrently with the *δ_o_* ≠ 0 CoVID-19 pandemic *Fast Shutoff*.

This analysis shows a wide variety of CoVID-19 data can be modeled using {*K_A_*, *γ_o_*, *δ_o_*, *t_offset_*} as parameters, covering: (I) an exponential rise at CoVID- 19 pandemic start; (II) a gradual lengthening of *doubling times* for a pandemic *Slow Shutoff*; and (III) an exponential decay for pandemic *Fast Shutoffs*.

## Data Availability

All data used is in the public domain or was on the CoVID-19 website maintained by bing.com.

